# National Population-Level Disparities in COVID-19 Mortality Across the Intersection of Race/Ethnicity and Sex in the United States

**DOI:** 10.1101/2021.08.29.21262775

**Authors:** Jay J. Xu, Jarvis T. Chen, Thomas R. Belin, Ronald S. Brookmeyer, Marc A. Suchard, Christina M. Ramirez

## Abstract

Males and certain racial/ethnic minority groups have borne a disproportionate burden of COVID-19 mortality in the United States, and substantial scientific research has sought to quantify and characterize population-level disparities in COVID-19 mortality outcomes by sex and across categories of race/ethnicity. However, there has not yet been a national population-level study to quantify disparities in COVID-19 mortality outcomes across the intersection of these demographic dimensions. Here, we analyze a publicly available dataset from the National Center for Health Statistics comprising COVID-19 death counts stratified by race/ethnicity, sex, and age for the year 2020, calculating mortality rates for each race/ethnicity-sex-age stratum and age-adjusted mortality rates for each race/ethnicity-sex stratum, quantifying disparities in terms of mortality rate ratios and rate differences. Our results reveal persistently higher COVID-19 age-adjusted mortality rates for males compared to females within every racial/ethnic group, with notable variation in the magnitudes of the sex disparity by race/ethnicity. However, non-Hispanic Black, Hispanic, and non-Hispanic American Indian or Alaska Native females have higher age-adjusted mortality rates than non-Hispanic White and non-Hispanic Asian/Pacific Islander males. Moreover, persistent racial/ethnic disparities are observed among both males and females, with higher COVID-19 age-adjusted mortality rates observed for non-Hispanic Blacks, Hispanics, and non-Hispanic American Indian or Alaska Natives relative to non-Hispanic Whites.

## 1 INTRODUCTION

The United States (U.S.) coronavirus disease 2019 (COVID-19) epidemic has impacted certain segments of the U.S. population disproportionately. For example, certain racial/ethnic minorities (Blacks, Hispanics, and American Indian or Alaska Natives) have disproportionately high numbers of COVID-19 cases, hospitalizations, and deaths (1). Men are also at disproportionately high risk of COVID-19-related hospitalization and mortality (2). We focus the attention of this paper on quantifying disparities in population-level COVID-19 mortality. Substantial scientific research has uncovered disparities in COVID-19 mortality outcomes in the U.S. by race/ethnicity (e.g., (3, 4, 5, 6, 7, 8, 9, 10, 11)), as well as between males and females (e.g., (12, 13, 14, 15, 16, 17, 18, 19)). However, research on U.S. population-level disparities in COVID-19 mortality outcomes across the intersection of race/ethnicity and sex has been limited, largely due to the absence of requisite publicly available data, which represents a critical research gap in the quantification and understanding of the nature of population-level disparities in COVID-19 mortality outcomes between subgroups.

The study by Rushovich et al. (2021) (20) was the first to investigate population-level disparities in COVID-19 mortality outcomes by race and sex jointly in the U.S. Their analysis centered around Michigan and Georgia, the only states that, at their time of writing, publicly provided the necessary stratified data to facilitate such analyses. Specifically, Rushovich et al. calculated COVID-19 mortality rates for each race-sex-age stratum, COVID-19 age-adjusted mortality rates for each race-sex stratum, and associated mortality rate ratios (RR) and rate differences (RD) to quantify disparities. They found that males had higher COVID-19 mortality rates than females within all racial groups considered, but the magnitudes of the sex disparity for different race categories varied substantially, illustrating the interactive nature of race and sex in their association with COVID-19 mortality outcomes.

Here, we extend the work of Rushovich et al. by quantifying such population-level disparities at the U.S. national level, leveraging available national data on COVID-19 deaths across racial/ethnic and sex categories. As such, our results are representative of the U.S. as a whole rather than just 2 states. Analogous to Rushovich et al., we calculate COVID-19 age-specific mortality rates for each race/ethnicity-sex-age stratum, COVID-19 age-adjusted mortality rates for each race/ethnicity-sex stratum, and associated mortality RR’s and RD’s. Our analysis includes a greater breadth of racial/ethnic groups, however. The analysis by Rushovich et al. did not account for Hispanic ethnicity and included only Whites, Blacks, and Asian/Pacific Islanders. In contrast, our analysis accounts for Hispanic ethnicity and includes additional racial groups: non-Hispanic Whites, non-Hispanic Blacks, Hispanics, non-Hispanic Asian or Pacific Islanders, and non-Hispanic American Indian or Alaska Natives. Lastly, our analysis improves upon the methodology employed in the Rushovich et al. study in the treatment of COVID-19 deaths with unknown race/ethnicity and in the handling of suppressed death counts. In their analysis, Rushovich et al. omitted COVID-19 deaths that had unknown race, and they also omitted small counts of COVID-19 deaths that were suppressed. This approach, however, underestimates the mortality rates and could potentially induce bias in the mortality RR and RD estimates. In contrast, we handle COVID-19 death records with missing race/ethnicity and/or suppressed death counts by calculating bounds for the possible mortality rate values consistent with the data, which subsequently determine bounds for the possible mortality RR and RD values. As such, we communicate the results of our analysis in terms of intervals prescribing the theoretically plausible range of the mortality rates, RR’s, and RD’s of interest in our analysis.

## 2 MATERIALS AND METHODS

### Data Sources

We examine a dataset from the National Center for Health Statistics (NCHS) that was publicly released on 15 April 2021, comprising COVID-19 death counts (ICD-10 code U07.1 (21) as an underlying or multiple cause of death) stratified by race/ethnicity, sex, and age for the year 2020 (22). Also included are counts of various underlying health conditions (e.g., diabetes mellitus, Alzheimer’s disease, influenza and pneumonia) of the COVID-19 decedents in the dataset, but we do not consider these factors in this paper. The sex categories are male and female, and the following 7 racial/ethnic groups are recorded: non-Hispanic White (NH White), non-Hispanic Black (NH Black), Hispanic, non-Hispanic Asian (NH Asian), non-Hispanic American Indian or Alaska Native (NH AIAN), non-Hispanic Native Hawaiian or Other Pacific Islander (NH NHOPI), and Unknown. The following set of age groups are used: 0–17, 18–29, 30–49, 50–64, and 65+. Death counts within race/ethnicity-sex-age strata between 1 and 9 are suppressed for confidentiality reasons to avoid the possibility that individuals would be identifiable. The NCHS dataset comprises 381,415 total non-suppressed COVID-19 deaths, but 1,620 of them have unknown race/ethnicity but known age and sex. Furthermore, there are 4 sex-age strata with unknown race/ethnicity that have suppressed death counts, and there are 6 sex-age strata with known race/ethnicity that have suppressed death counts. In this paper, we combine the NH Asian and NH NHOPI racial/ethnic groups into a single racial/ethnic group that we call non-Hispanic Asian or Pacific Islanders (NH Asian/PI) and combine their suppressed and non-suppressed death counts as appropriate. The NCHS data is included in the Supplementary Material as File S1.

To obtain the denominators for the mortality rates we calculate, we make use of estimated 2019 U.S. national population counts stratified by race/ethnicity, sex, and age from CDC WONDER (23). The racial/ethnic categories in the CDC WONDER data include the racial/ethnic groups we consider in our analysis, and the CDC WONDER age groups are integers from 0 to 84 and a catch-all 85+ age group. The estimated population counts are appropriately collapsed over age groups to conform to the age groups in the NCHS data. The CDC WONDER data is included in the Supplementary Material as File S2.

### Estimation Strategy for Handling Missing Race/Ethnicity and Suppressed Death Counts

As mentioned previously, 1,620 non-suppressed COVID-19 deaths in the NCHS data have unknown race/ethnicity, and 10 race/ethnicity-sex-age strata have suppressed death counts, each of which is an integer between 1 and 9. Our general estimation strategy is to calculate upper and lower bounds for the mortality rates of interest, which subsequently determine the upper and lower bounds for the associated mortality RR’s and RD’s. The upper bounds for the age-specific and age-adjusted mortality rates are obtained by (i) appropriately combining COVID-19 death counts with unknown race/ethnicity and COVID-19 death counts for the racial/ethnic group of interest, and (ii) assuming the maximal number (9) of deaths corresponding to suppressed death counts for the racial/ethnic group of interest or those with unknown race/ethnicity. The lower bounds for the age-specific and age-adjusted mortality rates are obtained by (i) assuming COVID-19 death counts with unknown race/ethnicity all do not correspond to the racial/ethnic group of interest, and (ii) assuming the minimal number (1) of deaths corresponding to suppressed death counts for the racial/ethnic group of interest. The upper and lower bounds for the age-specific and age-adjusted mortality rate ratios and differences are subsequently obtained from the appropriate quotients and differences of the upper and lower bounds for the age-specific and age-adjusted mortality rates. The mathematical details can be found in Appendix A. Hence, we communicate our results in terms of intervals prescribing the upper and lower bounds for the mortality rates, RR’s, and RD’s of interest.

### Computation

All calculations in our analysis are performed using the R programming language. The code used in our analysis is available upon reasonable request from the corresponding author.

## 3 RESULTS

Table 1 contains the theoretical bounds for the mortality rates for each race/ethnicity-sex-age stratum and the male-to-female mortality RR’s and RD’s for each race/ethnicity-age stratum. From Table 1, males have higher COVID-19 mortality rates than females within almost all race/ethnicity-age strata. For example, among individuals aged 50-64, the age-adjusted mortality rates among NH White men and women are (6.34–6.45) and (3.57–3.61) per 10,000, respectively; the age-adjusted mortality rates among NH Black men and women are (22.1–22.8) and (13.3–13.5) per 10,000, respectively; the age-adjusted mortality rates among Hispanic men and women are (30.2–30.7) and (12.4–12.6) per 10,000, respectively; the age-adjusted mortality rates among NH Asian/PI men and women are (10.4–11.8) and (3.33–3.74) per 10,000, respectively; and the age-adjusted mortality rates among NH AIAN men and women are (29.9–40.2) and (19.6–22.7) per 10,000, respectively.

**Table 1.** Theoretical bounds for the (i) mortality rates in each race/ethnicity-sex-age stratum, (ii) male-to-female mortality rate ratios for each race/ethnicity-age stratum, and (iii) male-to-female mortality rate differences for each race/ethnicity-age stratum. Quantities are calculated with respect to data on COVID-19 deaths from the National Center for Health Statistics for the year 2020. Mortality rates are expressed per 10,000 people.

| Race/Ethnicity | Age | Mortality Rate |  |  |  |
| --- | --- | --- | --- | --- | --- |
|  |  | Male | Female | Ratio | Difference |
| NH White | 0-17 years | (0.01–0.02) | (0.01–0.02) | (0.63–1.17) | (-0.01, 0.00) |
|  | 18-29 years | (0.13–0.14) | (0.09–0.09) | (1.43–1.60) | (0.04–0.05) |
|  | 30-49 years | (0.88–0.90) | (0.55–0.56) | (1.59–1.64) | (0.33–0.35) |
|  | 50-64 years | (6.34–6.45) | (3.57–3.61) | (1.76–1.81) | (2.73–2.88) |
|  | 65+ years | (55.6–56.0) | (44.7–44.9) | (1.24–1.25) | (10.7–11.3) |
| NH Black | 0-17 years | (0.06–0.07) | (0.03–0.05) | (1.07–2.04) | (0.00–0.04) |
|  | 18-29 years | (0.50–0.53) | (0.40–0.42) | (1.20–1.32) | (0.08–0.13) |
|  | 30-49 years | (4.06–4.13) | (2.62–2.65) | (1.53–1.58) | (1.41–1.51) |
|  | 50-64 years | (22.1–22.8) | (13.3–13.5) | (1.64–1.72) | (8.63–9.50) |
|  | 65+ years | (106.0–109.8) | (71.4–72.9) | (1.45–1.54) | (33.1–38.5) |
| Hispanic | 0-17 years | (0.04–0.05) | (0.03–0.04) | (1.04–1.64) | (0.00–0.02) |
|  | 18-29 years | (0.67–0.69) | (0.36–0.37) | (1.80–1.92) | (0.30–0.33) |
|  | 30-49 years | (5.69–5.74) | (2.18–2.20) | (2.59–2.64) | (3.49–3.56) |
|  | 50-64 years | (30.2–30.7) | (12.4–12.6) | (2.40–2.48) | (17.6–18.3) |
|  | 65+ years | (124.2–128.1) | (71.5–73.2) | (1.70–1.79) | (51.0–56.7) |
| NH Asian/PI | 0-17 years | (0.01–0.12) | (0.01–0.13) | (0.07–13.0) | (-0.12, 0.11) |
|  | 18-29 years | (0.21–0.26) | (0.09–0.18) | (1.18–2.98) | (0.03–0.17) |
|  | 30-49 years | (1.50–1.64) | (0.65–0.70) | (2.15–2.52) | (0.80–0.99) |
|  | 50-64 years | (10.4–11.8) | (3.33–3.74) | (2.77–3.56) | (6.63–8.50) |
|  | 65+ years | (53.5–60.4) | (32.8–35.9) | (1.49–1.84) | (17.6–27.6) |
| NH AIAN | 0-17 years | (0.03–0.49) | (0.00–0.25) | (0.11, +∞) | (-0.22, 0.49) |
|  | 18-29 years | (1.47–1.81) | (0.93–1.28) | (1.15–1.95) | (0.19–0.88) |
|  | 30-49 years | (9.22–10.5) | (6.71–7.20) | (1.28–1.56) | (2.02–3.73) |
|  | 50-64 years | (29.9–40.2) | (19.6–22.7) | (1.32–2.05) | (7.28–20.6) |
|  | 65+ years | (95.0–148.2) | (71.9–96.4) | (0.99–2.06) | (-1.38, 76.3) |

Table 2 presents the theoretical bounds for the COVID-19 age-adjusted mortality rates for males and females stratified by race/ethnicity along with their corresponding age-adjusted male-to-female mortality RR’s and RD’s. To complement Table 2, Figure 1 graphically illustrates the male and female age-adjusted mortality rates stratified by race/ethnicity, and Figure 1 graphically illustrates the age-adjusted male-to-female mortality RR’s and RD’s stratified by race/ethnicity. Males have higher age-adjusted mortality rates than females within each racial/ethnic group considered, with some differences quite notable. On the ratio scale, the age-adjusted mortality rate for males is over 60% higher than that of females among NH Asian/PI’s and over 80% higher among Hispanics. On the difference scale, the male-female differences in age-adjusted mortality rates are highest among Hispanics (12.7–13.8 deaths per 10,000) and NH Blacks (7.48–8.58 deaths per 10,000).

**Table 2.** Theoretical bounds for the (i) age-adjusted mortality rates for each race/ethnicity-sex stratum, (ii) age-adjusted male-to-female mortality rate ratios stratified by race/ethnicity, and (iii) age-adjusted male-to-female mortality rate differences stratified by race/ethnicity. Quantities are calculated with respect to data on COVID-19 deaths from the National Center for Health Statistics for the year 2020. Mortality rates are expressed per 10,000 people.

| Race/Ethnicity | Age-Adjusted Mortality Rate |  |  |  |
| --- | --- | --- | --- | --- |
|  | Male | Female | Ratio | Difference |
| NH White | (10.6–10.7) | (8.20–8.25) | (1.29–1.31) | (2.37–2.51) |
| NH Black | (22.8–23.6) | (15.0–15.3) | (1.49–1.57) | (7.48–8.58) |
| Hispanic | (27.8–28.6) | (14.8–15.1) | (1.84–1.94) | (12.7–13.8) |
| NH Asian/PI | (11.2–12.7) | (6.23–6.86) | (1.64–2.04) | (4.36–6.48) |
| NH AIAN | (24.0–35.2) | (17.5–22.3) | (1.07–2.02) | (1.67–17.7) |

**Figure 1.**
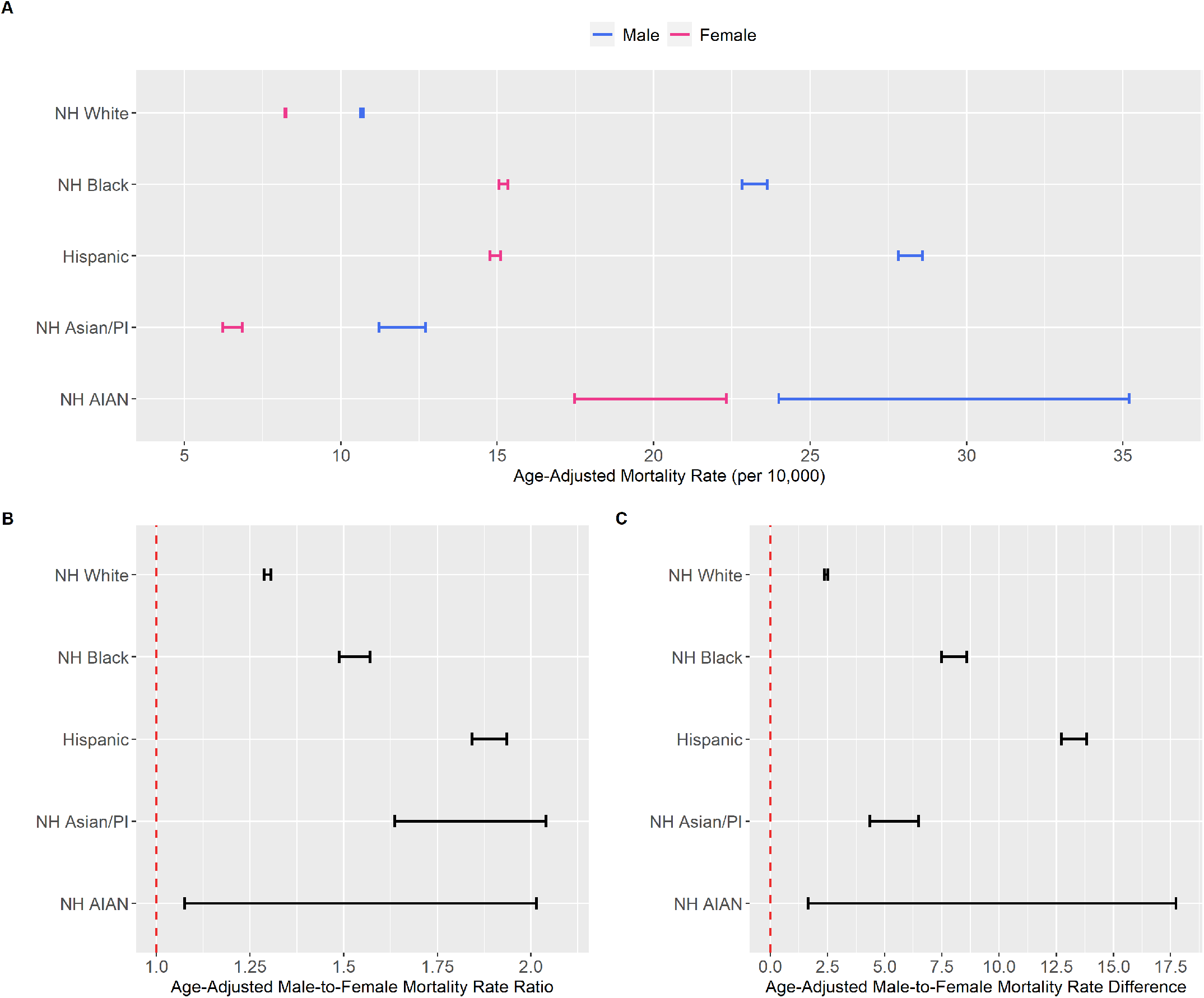
**(A)** Theoretical bounds for the age-adjusted mortality rates for each race/ethnicity-sex stratum, **(B)** Theoretical bounds for the age-adjusted male-to-female mortality rate ratios by race/ethnicity, **(C)** Theoretical bounds for the age-adjusted male-to-female mortality rate differences by race/ethnicity. Quantities calculated from data on COVID-19 deaths from the National Center for Health Statistics for the year 2020. Mortality rate differences are expressed per 10,000 people.

The notable variation in the magnitudes of the sex disparity across different racial/ethnic categories is consistent with the findings of Rushovich et al. The male-female disparity is greater among NH Blacks and NH Asian/PI’s than among NH Whites on both the ratio and difference scales, and the male-female disparity is greater for Hispanics than both NH Whites and NH Blacks on both the ratio and difference scales. The male-female disparity is greater among NH Asian/PI’s than among NH Blacks on the ratio scale, but the reverse is true on the difference scale. While the level of detail in the NCHS data was sufficient to allow us to ascertain that the COVID-19 age-adjusted mortality rate for males is greater than that for females among NH AIAN’s, the widths of the corresponding age-adjusted male-to-female mortality RR and RD intervals obscures comparisons of the magnitude of the male-female disparity between NH AIAN’s and the other racial/ethnic groups.

A second way to characterize the disparities is to compare racial/ethnic groups within sex categories. To this end, Table 3 presents the theoretical bounds for the COVID-19 age-adjusted mortality rates for NH Whites, NH Blacks, Hispanics, NH Asian/PI’s, and NH AIAN’s stratified by sex along with their corresponding age-adjusted mortality RR’s and RD’s, anchoring comparisons to NH Whites. From Table 3, the magnitudes of the racial/ethnic disparities in COVID-19 mortality are often substantial but vary by sex. Disparities between NH Blacks and Hispanics relative to NH Whites are greater among males than among females on both the ratio and difference scales. Among males, age-adjusted mortality rates for NH Blacks are over twice that of NH Whites, and age-adjusted mortality rates for Hispanics are over 2.5 times that of NH Whites. Among females, age-adjusted mortality rates for NH Blacks and Hispanics are over 75% higher than that for NH Whites. Remarkably, age-adjusted mortality rates for NH AIAN’s are over twice that of NH Whites among both males and females. Among males, the COVID-19 age-adjusted mortality rate is lowest among NH Whites, followed by NH Asian/PI’s. Among females, however, the order is switched: the COVID-19 age-adjusted mortality rate is lowest among NH Asian/PI’s, followed by NH Whites. Among males, the COVID-19 age-adjusted mortality rate for Hispanics is notably greater than that of NH Blacks, but the COVID-19 age-adjusted mortality rates between NH Blacks and Hispanics are indistinguishable (overlapping intervals) among females. Among females, the COVID-19 age-adjusted mortality rate is highest among NH AIAN’s, but among males, the COVID-19 age-adjusted mortality rate for NH AIAN’s is higher than those of NH Whites, NH Blacks, and NH Asian/PI’s but is indistinguishable from that of Hispanics. Interestingly, COVID-19 age-adjusted mortality rates are higher for NH Asian/PI’s than NH Whites among males but lower among females.

**Table 3.** Theoretical bounds for the (i) age-adjusted mortality rates for each race/ethnicity-sex stratum; (ii) age-adjusted mortality rate ratios for NH Blacks, Hispanics, NH Asian/PI’s, and NH AIAN’s relative to NH Whites, stratified by sex; and (iii) age-adjusted mortality rate differences for NH Blacks, Hispanics, NH Asian/PI’s, and NH AIAN’s relative to NH Whites, stratified by sex. Quantities are calculated with respect to data on COVID-19 deaths from the National Center for Health Statistics for the year 2020. Mortality rates are expressed per 10,000 people.

| Sex | Race/Ethnicity | Age-Adjusted Mortality Rate |  |  |
| --- | --- | --- | --- | --- |
|  |  | Rate | Ratio | Difference |
| Male | NH White | (10.6–10.7) | Reference | Reference |
|  | NH Black | (22.8–23.6) | (2.13–2.22) | (12.1–13.0) |
|  | Hispanic | (27.8–28.6) | (2.60–2.69) | (17.1–18.0) |
|  | NH Asian/PI | (11.2–12.7) | (1.05–1.20) | (0.50–2.09) |
|  | NH AIAN | (24.0–35.2) | (2.24–3.31) | (13.3–24.6) |
| Female | NH White | (8.20–8.25) | Reference | Reference |
|  | NH Black | (15.0–15.3) | (1.82–1.87) | (6.80–7.14) |
|  | Hispanic | (14.8–15.1) | (1.79–1.84) | (6.52–6.90) |
|  | NH Asian/PI | (6.23–6.86) | (0.76–0.84) | (-2.02, -1.35) |
|  | NH AIAN | (17.5–22.3) | (2.12–2.72) | (9.22–14.1) |

Stark contrasts in COVID-19 age-adjusted mortality rates are also observed between certain subgroups defined by race/ethnicity and sex. For example, COVID-19 age-adjusted mortality rates among Hispanic males are over 3 times that of NH White females and over 4 times that of NH Asian/PI females. Furthermore, although COVID-19 age-adjusted mortality rates are higher among males within all racial/ethnic groups, females among some racial/ethnic groups have greater COVID-19 age-adjusted mortality rates than males in other racial/ethnic groups: NH Black, Hispanic, and NH AIAN females all have higher COVID-19 age-adjusted mortality rates than NH White and NH Asian/PI males, important details that are masked when analyzing COVID-19 mortality data disaggregated by sex but not also by race/ethnicity. Indeed, the age-adjusted mortality rates for both NH Black and Hispanic females are over 40% higher than that of NH White males, and the NH AIAN female age-adjusted mortality rate is over 60% higher than that of NH White males.

## 4 DISCUSSION

This is the first analysis quantifying population-level disparities in COVID-19 mortality across race/ethnicity and sex jointly at the U.S. national level. Having demonstrated empirically that males have higher COVID-19 age-adjusted mortality rates within each racial/ethnic group, certain racial/ethnic minority groups (non-Hispanic Blacks, Hispanics, and non-Hispanic AIAN’s) have higher COVID-19 age-adjusted mortality rates relative to non-Hispanic Whites among both males and females, and sex differences in COVID-19 age-adjusted mortality rates vary markedly in magnitude by race/ethnicity, researchers must consider multiple pathways (e.g., biological, behavioral, social), that could be responsible for driving these observed differences in COVID-19 mortality outcomes across the intersection of race/ethnicity and sex.

The persistent one-sided and male-biased disparity within racial/ethnic groups lends support to the theory that biological differences between males and females play an important role in driving the pronounced differences in COVID-19 mortality rates by sex (24, 25, 26, 27). Indeed, there has been extensive scientific research in this domain. An abundance of attention has been focused on differences in angiotensin-converting enzyme-2 (ACE2) (28, 29, 30) expressions, transmembrane serine protease 2 (TMPRSS2) (31, 32) expressions, and T-cell response (33, 34, 35) between males and females and their relationship to observed disparities in COVID-19 mortality outcomes by sex. Differential rates of certain pre-existing health conditions by sex have also been cited as a likely contributing factor to the differential rates of COVID-19 mortality by sex. For instance, U.S. men have higher rates of coronary artery disease (36, 37), diabetes (38), and liver disease (38), which are risk factors for COVID-19-related severe outcomes (39). Men in the U.S. also have higher rates of hypertension (40), which some studies have indicated may also be a risk factor for COVID-19-related severe outcomes (39, 41, 42).

Behavioral differences between men and women have also been widely hypothesized as a contributing factor to the observed male-female disparities in COVID-19 mortality. For example, men have higher rates of cigarette smoking (43), which is a risk factor for COVID-19-related severe outcomes (39). Men are less likely to wear face masks than women (44, 45, 46, 47, 48), and some studies have suggested that masks can reduce the severity of a COVID-19 infection by reducing the viral inoculum (49, 50, 51, 52). Proper hand washing may also reduce the viral inoculum of a COVID-19 infection (53), and men are much less likely to practice proper hand washing behaviors (54, 55, 56). More broadly, women have been estimated to be approximately 50% more likely than men to practice non-pharmaceutical health-protective behaviors (e.g., proper hand washing, face mask wearing, surface cleaning) in the context of respiratory infectious disease epidemic and pandemic settings, according to a 2016 meta-analysis (57).

Our results also demonstrated that NH Blacks, Hispanics, and NH AIAN’s have substantially higher COVID-19 age-adjusted mortality rates compared to NH Whites among both males and females, which is concerning from a health equity perspective. The disproportionately high COVID-19 mortality burden experienced by these racial/ethnic groups is well-documented, and many theories have been proposed to explain these disparities. For example, differences in the rates by race/ethnicity of pre-existing health conditions identified as risk factors for severe COVID-19 illness (39) have been theorized as contributing to observed disparities in COVID-19 mortality outcomes by race/ethnicity. Blacks have substantially higher rates of diabetes and hypertension relative to NH Whites (58, 59), as well as an overall higher rate of obesity (60). Hispanics have a considerably higher rate of diabetes than NH Whites (61) as well as a higher overall rate of obesity (62). NH AIAN’s have nearly three times the rate of diabetes as NH Whites (63) as well as higher overall rates of coronary heart disease and hypertension (64). Health insurance coverage rates among NH Blacks, Hispanics, and NH AIAN’s have historically lagged behind NH Whites and NH Asians (65), which generally contributes to poorer health outcomes and higher rates of chronic health conditions, some of which are listed above. In fact, a Gallup-West Health study conducted early in the U.S. COVID-19 epidemic (1–14 April 2020) found that 14% of non-White survey respondents would avoid treatment for a suspected COVID-19 infection due to cost of care compared to only 6% of Whites (66). Poor housing conditions, inextricably linked with overcrowding and lack of adequate plumbing and sanitation, have been found to be associated with higher COVID-19 incidence and mortality (67), and NH Blacks and Hispanics are more likely to reside in moderate to severe substandard housing than NH Whites (68). Blacks and Hispanics are also more likely to live in areas with higher rates of industrial air pollution (69), which some studies have suggested may increase the risk of COVID-19 mortality (70, 71, 72).

The notable variation in the magnitudes of the male-female disparities across race/ethnicity (or equivalently, the notable variation in the magnitudes of the racial/ethnic disparities across sex) indicates that factors related to social determinants of health (73, 74)—some examples of which are listed above—play an important role in driving disparities in COVID-19 mortality outcomes across population subgroups defined by race/ethnicity and sex. For example, the sex disparity in the COVID-19 age-adjusted mortality rates among Hispanics is notably larger than those among NH Whites and NH Blacks in terms of both age-adjusted mortality RR’s and RD’s. A possible reason for such heterogeneity is the fact that Hispanic men are vastly overrepresented in essential industries such as farming/agriculture, construction, and transportation (75, 76) that required continued employment during the COVID-19 pandemic, subjecting them to increased risks of COVID-19 exposure. Indeed, a 2021 study in California (U.S.) indicated that these industries are among the occupational sectors with the highest associated excess mortality attributable to COVID-19 (77). Our results are consistent with this hypothesis, with markedly greater male-to-female mortality RR’s and RD’s observed among Hispanics compared to NH Blacks and NH Whites in the prime working age groups (Table 1). Moreover, even though COVID-19 age-adjusted mortality rates are higher for males compared to females within every racial/ethnic group, this pattern does not hold across all racial/ethnic groups. The fact that NH Black, Hispanic, and NH AIAN females all have higher COVID-19 age-adjusted mortality rates than NH White and NH Asian/PI males invalidates the potential view that observed disparities by sex are entirely or mostly a consequence of innate biological differences that are constant across racial/ethnic groups. Indeed, the perspective that factors related to social determinants of health primarily drive the racial/ethnic disparities in COVID-19 mortality outcomes themselves is widely propounded in the scientific community (78, 79, 80, 81, 82, 83). Nevertheless, precisely how biological factors and social processes both individually and jointly influence the nature and extent of population-level disparities in COVID-19 mortality outcomes across the intersection of race/ethnicity and sex is a complex scientific problem that warrants further research, but a comprehensive investigation is beyond the scope of this paper.

Finally, population-level disparities research is important in order to identify preventable morbidity and mortality and improve our knowledge of factors that contribute to disparities in health outcomes between population subgroups, with the ultimate hope of discovering interventions that will reduce them (84). This study highlights some of the limitations of previous analyses quantifying population-level disparities that relied on public datasets reporting limited socio-demographic characteristics associated with COVID-19 outcomes. Such low-dimensional data inherently limits our ability to understand of the nature of disparities in COVID-19 outcomes between population subgroups and does little to inform how such disparities might be reduced or eliminated. Sophisticated disparities research is only possible with COVID-19 data disaggregated by multiple key variables, but such publicly available datasets in the U.S. have been and continue to be limited. Examples of stratifying variables of practical interest include but are not limited to race/ethnicity, sex, age, educational attainment, occupation, income, religious affiliation, household composition, underlying medical conditions, and geographical residence. We urge U.S. federal, state, and local governmental and health authorities to prioritize the public availability of accurate, detailed, and real-time multi-dimensional data on socially-relevant patient/decedent characteristics associated with COVID-19 outcomes to facilitate research illuminating health disparities between population subgroups so that they can be addressed.

## Data Availability

The datasets analyzed for this study can be found in the Supplementary Material, available for download in the online version of the article.

## AUTHOR CONTRIBUTIONS

JJX contributed to the conceptualization of the project, development of the methodology, data analysis, visualization, and writing the original draft of the manuscript. JJX, JTC, TRB, RSB, MAS, and CMR contributed to reviewing and editing of the manuscript.

## DATA AVAILABILITY STATEMENT

### APPENDIX A

#### Age-Specific Mortality Rates by Race/Ethnicity and Sex

The upper bound for the age-specific mortality rate for race/ethnicity *r*, sex *s*, and age *a* is obtained using the following equation:

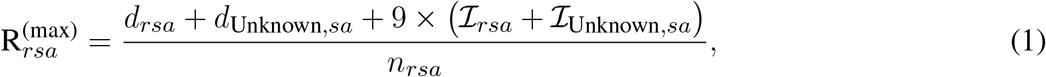

where *d*_*rsa*_ is the number of non-suppressed COVID-19 deaths corresponding to race/ethnicity *r*, sex *s*, and age group *a*; *ℐ*_*rsa*_ is the number of suppressed death counts corresponding to race/ethnicity *r*, sex *s*, and age group *a*; and *n*_*rsa*_ is the CDC WONDER 2019 U.S. national population estimate corresponding to race/ethnicity *r*, sex *s*, and age group *a*. Note that if *ℐ*_*rsa*_ *>* 0, then *d*_*rsa*_ = 0 except in the case of NH Asian/PI’s, where *d*_*rsa*_ can be greater than 0 because we combine the NH Asian and NH NHOPI racial/ethnic groups in our analysis. Also, note that *ℐ*_*rsa*_ can be only be 0 or 1 except in the case of NH Asian/PI’s, where it can also attain the value of 2 because we combine the NH Asian and NH NHOPI racial/ethnic groups in our analysis.

The lower bound for the age-specific mortality rate corresponding to race/ethnicity *r*, sex *s*, and age *a* is obtained using the following equation:

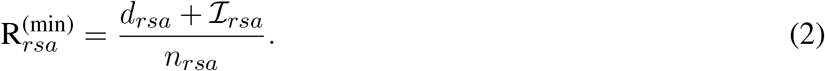

#### Age-Adjusted Mortality Rates by Race/Ethnicity and Sex

The age-adjusted mortality rates by race/ethnicity and sex are calculated using direct age adjustment (85). The upper bound for the age-adjusted mortality rate for race/ethnicity *r* and sex *s* is obtained using the following equation:

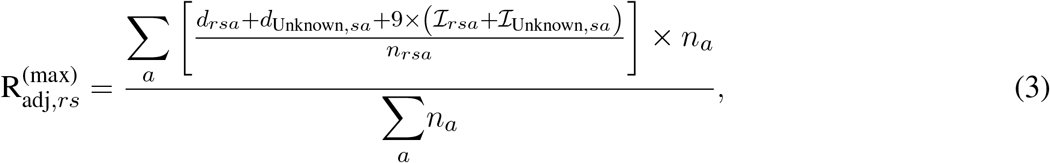

where *n*_*a*_ is the CDC WONDER 2019 U.S. national population estimate corresponding to age group *a*.

The lower bound for the age-adjusted mortality rate corresponding to race/ethnicity *r* and sex *s* is obtained using the following equation:

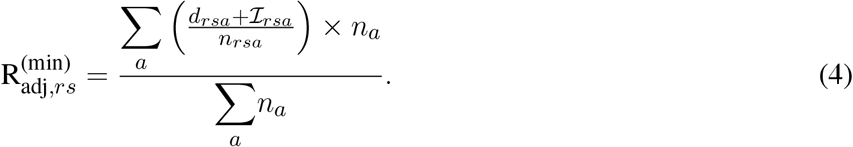

#### Age-Specific and Age-Adjusted Mortality Rate Ratios and Mortality Rate Differences

Theoretical bounds for the age-specific and age-adjusted mortality RR’s and RD’s can be obtained in a straightforward manner from the appropriate upper and lower bounds for the corresponding individual mortality rates.

To obtain the theoretical bounds for the age-specific mortality RR/RD comparing race/ethnicity-sex-age stratum (*r, s, a*) to stratum 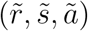 (reference stratum), the upper bound equals the upper bound of the mortality rate for stratum (*r, s, a*), given in Equation (1), divided by/minus the lower bound of the mortality rate for stratum 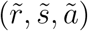, given in Equation (2):

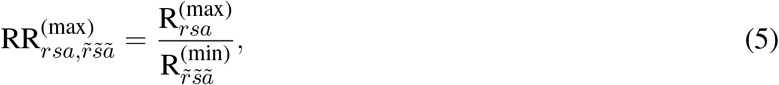

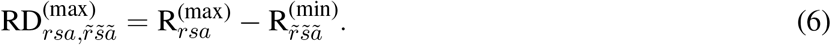

The lower bound of the age-specific mortality RR/RD comparing race/ethnicity-sex-age stratum (*r, s, a*) to stratum 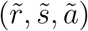 equals the lower bound of the mortality rate for stratum (*r, s, a*) divided by/minus the upper bound of the mortality rate for stratum 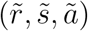:

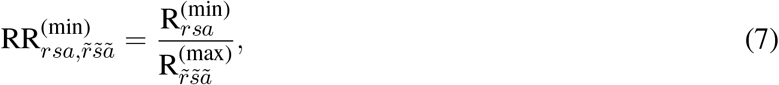

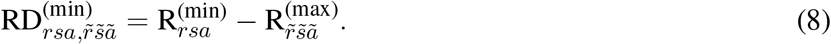

The upper bound of the age-adjusted mortality RR/RD comparing race/ethnicity-sex stratum (*r, s*) to stratum 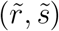 (reference stratum) equals the upper bound of the age-adjusted mortality rate for stratum (*r, s*), given in Equation (3), divided by/minus the lower bound of the age-adjusted mortality rate for stratum 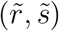, given in Equation (4):

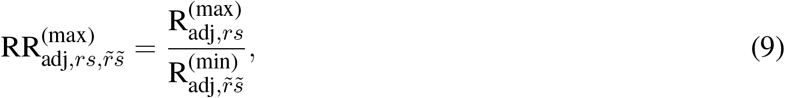

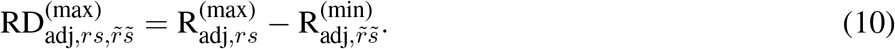

The lower bound of the age-adjusted mortality RR/RD comparing race/ethnicity-sex stratum (*r, s*) to stratum 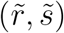 equals the lower bound of the age-adjusted mortality rate for stratum (*r, s*) divided by/minus the upper bound of the age-adjusted mortality rate for stratum 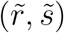:

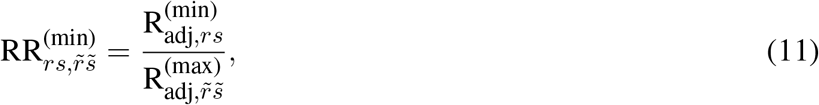

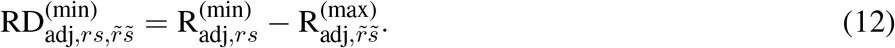

